# Moving beyond self-report in characterizing drug addiction: Using drug-biased behavior to prospectively inform treatment adherence in opioid use disorder

**DOI:** 10.1101/2025.01.01.25319860

**Authors:** Natalie McClain, Ahmet O. Ceceli, Greg Kronberg, Nelly Alia-Klein, Rita Z. Goldstein

## Abstract

Drug addiction is accompanied by enhanced salience attributed to drug over nondrug cues. This objectively measured bias is reliable yet underutilized in informing clinical endpoints, as clinical trials largely employ subjective (i.e., self-report or interview-based drug use and craving) or simple categorical (e.g., drug in urine) measures, with limited success. Having previously demonstrated their utility in cocaine addiction, we investigated whether behavioral picture choice (a lab-simulated drug seeking measure) and verbal fluency similarly reveal drug bias in 59 abstinent, inpatient individuals with opioid use disorder (iOUD) compared to 29 healthy controls (HC). Using a hierarchical regression, and compared to subjective measures, we then tested whether these objective markers can better inform prospective treatment completion—a clinically relevant and measurable outcome. As expected, results showed that the iOUD exhibited higher simulated drug seeking (*p*s*<*0.036) and drug fluency (*p=*0.008) compared to the HC. Importantly, after dimensionality reduction, while the self-reported years of regular opioid use and cue-induced craving showed null results (|*β*|<0.47, *p*>0.290), and controlling for demographics, drug choice was associated with treatment completion *β* =-0.75, *p*=0.036), explaining greater variability in its likelihood compared to the subjective measures (model comparison: *ΔR*^*2*^=0.102, *p*=0.027). Extending drug-biased choice and fluency from cocaine to opioid addiction, results further indicate that these objective measures of drug bias outperform the commonly employed subjective drug use and craving in informing a clinical outcome; unlike drug urine tests, they show important variability in abstinent iOUD. Results implicate these cognitive-behavioral tasks as powerful markers of drug bias and predictors of treatment outcome.

## 1. INTRODUCTION

Drug addiction continues to pose a grave public health concern, evidenced by the rising number of overdose deaths, exceeding 100,000 in 2022 and largely driven by opioids, including heroin and fentanyl co-use [1,2]. Accompanying the chronically relapsing nature of drug addiction [3] is a high risk of premature treatment dropout, with rates averaging 30% across drugs of abuse, with estimates as high as 91% in opioid addiction [4–6]. Thus, identifying markers of treatment retention is imperative for relapse vulnerability detection and its putative prevention. According to the impaired response inhibition and salience attribution (iRISA) model, a core symptom that perpetuates drug addiction and deters successful recovery is the attribution of excessive value to drug cues at the expense of nondrug cues and reinforcers [7,8]. Measures of this drug bias could reliably characterize addiction severity, potentially serving as robust objective predictors of clinical outcomes.

Clinical trials that target addiction treatment have relied heavily on subjective (i.e., self-report and/or interview-based) measures to ascertain addiction severity and treatment endpoints. These self-report measures are useful in assessing retrospective patterns of use [9], readiness for change and motivations to use [10], and living conditions and support systems [11] to construct a comprehensive addiction profile. Self-reported craving scales have also been useful in predicting proximal use [12], relapse [13], and treatment dropout [14], especially in research settings with time or resource constraints [15]. However, self-reported (declarative) measures are susceptible to reporting errors, inclusive of a more biased recall (e.g., under-reporting) of distant vs. proximal events ([16–18]; but also see [19]), and can be influenced by compromises in self-awareness and insight into illness [20–22], demand characteristics [23], and social desirability [24]. For example, a study comparing self-report with actual clinic records in drug treatment patients found heightened misreporting of heroin use [25], potentially due to greater social stigma [25,26]. The accuracy of self-report may also be sensitive to treatment stage, as suggested by increased disagreement between self-reported drug use and urine toxicology as a function of time in treatment in outpatient iOUD [27], together limiting the applicability of self-report in long-term treatment environments. Clinical research to evaluate outcomes has also used urine toxicology, yet it is a binary measure insufficient for capturing dynamic changes in drug use patterns [28] and is non-variable in certain contexts (e.g., drug urine-negative results in controlled treatment or correctional settings). Together, additional approaches are needed to better characterize clinical endpoints in drug addiction treatment—and especially for opioid addiction, affecting the majority of those seeking treatment for (non-alcohol) drug use [29,30].

The literature on the utility of objective cognitive-behavioral measures to inform treatment outcomes in addiction is scarce (see for a viewpoint [31]). Nevertheless, neuropsychological and behavioral measures of impulsivity [32–34], working memory [34], decision-making [35], and other forms of cognitive functioning (attention, visual-verbal memory, visuospatial ability) [36] predict treatment dropout/retention. For example, a behavioral risky decision-making task assessing ambiguity tolerance (pursuing conditions where outcome odds are unknown) predicted prospective use in iOUD [37]. While these tasks are informative, it is essential to tailor these measures to drug-related cues/contexts to tap into the core impairments and underlying neural substrates in drug addiction [7,8]. For example, behavioral measures of drug-biased attention (e.g., modified Stroop and visual probe tasks) predict clinical outcomes ([38,39]; in heroin addiction, see [40,41]; in opioid dependence see [42,43]). We demonstrated that an electroencephalographic measure of motivated drug-biased attention captured incubation of drug cue-induced reactivity in cocaine addiction, whereas subjective ratings of baseline craving did not [44], highlighting the former as a more sensitive marker of cue-induced relapse vulnerability. Targeting higher-order executive functions, we further showed that cocaine-biased (cocaine>pleasant) picture choice (a lab-simulated measure of drug seeking) was associated with recent and prospective (6-months) use in cocaine addiction [21,45,46]. Linguistic markers from speech about drug-use consequences predicted longitudinal outcomes (craving, withdrawal, abstinence, drug use), outperforming non-linguistic models (using demographics, neuropsychological, and drug-use measures) in predicting 12-months abstinence in cocaine addiction [47]. Taken together, serving as sensitive and reliable proxies of drug-use severity, and compared to more neutral measures or self-reports, cognitive-behavioral tasks that target drug-biased processing may better predict clinical outcomes in drug addiction.

Here we tested whether select cognitive-behavioral markers of drug bias outperform demographics and other traditional measures of addiction severity in informing prospective (4-months) treatment adherence in inpatient iOUD. The markers included 1) picture-viewing choice tasks (developed in cocaine addiction [45,46], then validated across other drugs [48–51]) and 2) a verbal fluency task designed to probe attentional bias and conditioned reactivity towards drug-related words in cocaine addiction [52,53]. Attendance of a follow-up study visit, after completing a randomized treatment component within a longitudinal study, served as our objective clinical endpoint, previously used for this purpose [54,55] including in clinical trials [56,57]. This measure shows a strong relationship with post-treatment success [58–60] and bears relevance across treatment types (e.g., inpatient residential, outpatient). We hypothesized that 1) the iOUD would show higher drug-biased choice and fluency compared to HC and 2) these objective drug-bias measures would outperform demographics, subjective drug-use and craving measures in informing treatment completion. As an exploratory hypothesis, we anticipated reductions in drug-biased behavior at follow-up in iOUD.

## 2. METHODS

### 2.1. Participants

Fifty-nine iOUD from a rehabilitation facility and 29 age- and sex-matched HC from the surrounding community participated in this study (see Table 1). Participants provided informed consent in accordance with the Icahn School of Medicine at Mount Sinai’s Institutional Review Board. All iOUD were inpatients at a residential drug treatment program, receiving relapse prevention treatment, anger management training, Seeking Safety therapy (a present-focused, cognitive-behavioral therapy model targeting trauma and/or substance abuse), and other counseling services. All iOUD were abstinent (160.58 ± 188.18 days) and stabilized on methadone (n=52; 112.94 ± 60.75 mg) or suboxone (n=7; 13.33 ± 8.64) at baseline. A comprehensive diagnostic interview, encompassing the Mini International Neuropsychiatric Interview 7^th^ ed. [61] and the Addiction Severity Index 5^th^ ed. [62], was performed to assess DSM-5 criteria for major psychiatric and substance use disorders. All iOUD met criteria for OUD with heroin as their primary substance/reason for treatment. See the supplement for details on eligibility criteria and psychiatric comorbidities. Heroin dependence, withdrawal, and craving were evaluated via the Severity of Dependence Scale [63], the Subjective Opiate Withdrawal Scale [64], and the Heroin Craving Questionnaire (modified from Cocaine Craving Questionnaire [65]), respectively. Nicotine dependence was measured with the Fagerström Test for Nicotine Dependence [66]. Picture cue- and movie scene-craving scores were collected via self-reported ratings of picture stimuli from an in-house drug cue reactivity task [67] and clips from a drug-related movie (3-sec clips sampled every 30 seconds from the first 17-minutes of the movie *Trainspotting* [68]), respectively. Cue-related drug wanting>liking (of the last drug use to intoxication) was collected via the Sensitivity To Reinforcement of Addictive and Other Primary Rewards (STRAP-R) questionnaire [69]. Depression and anxiety severity were measured using Beck’s Depression [70] and Anxiety [71] Inventories, respectively.

**Table 1.**
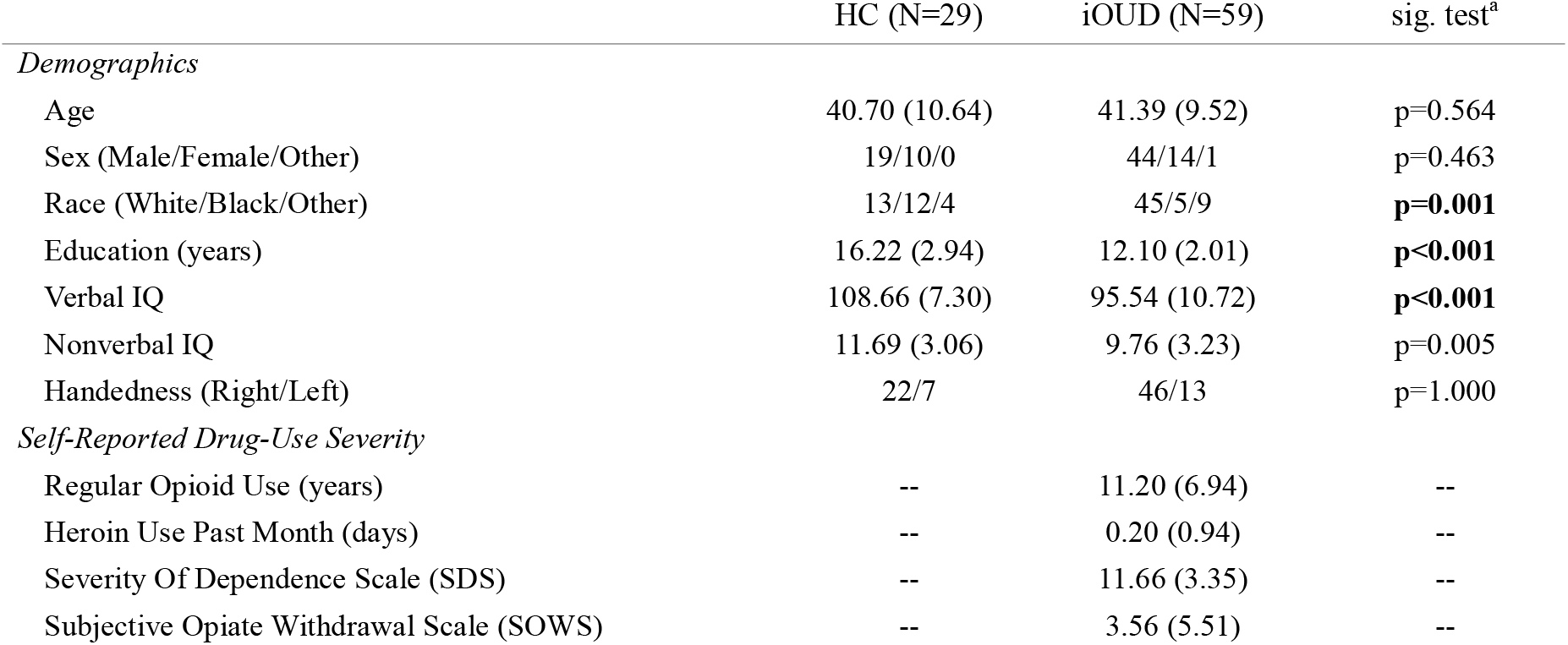
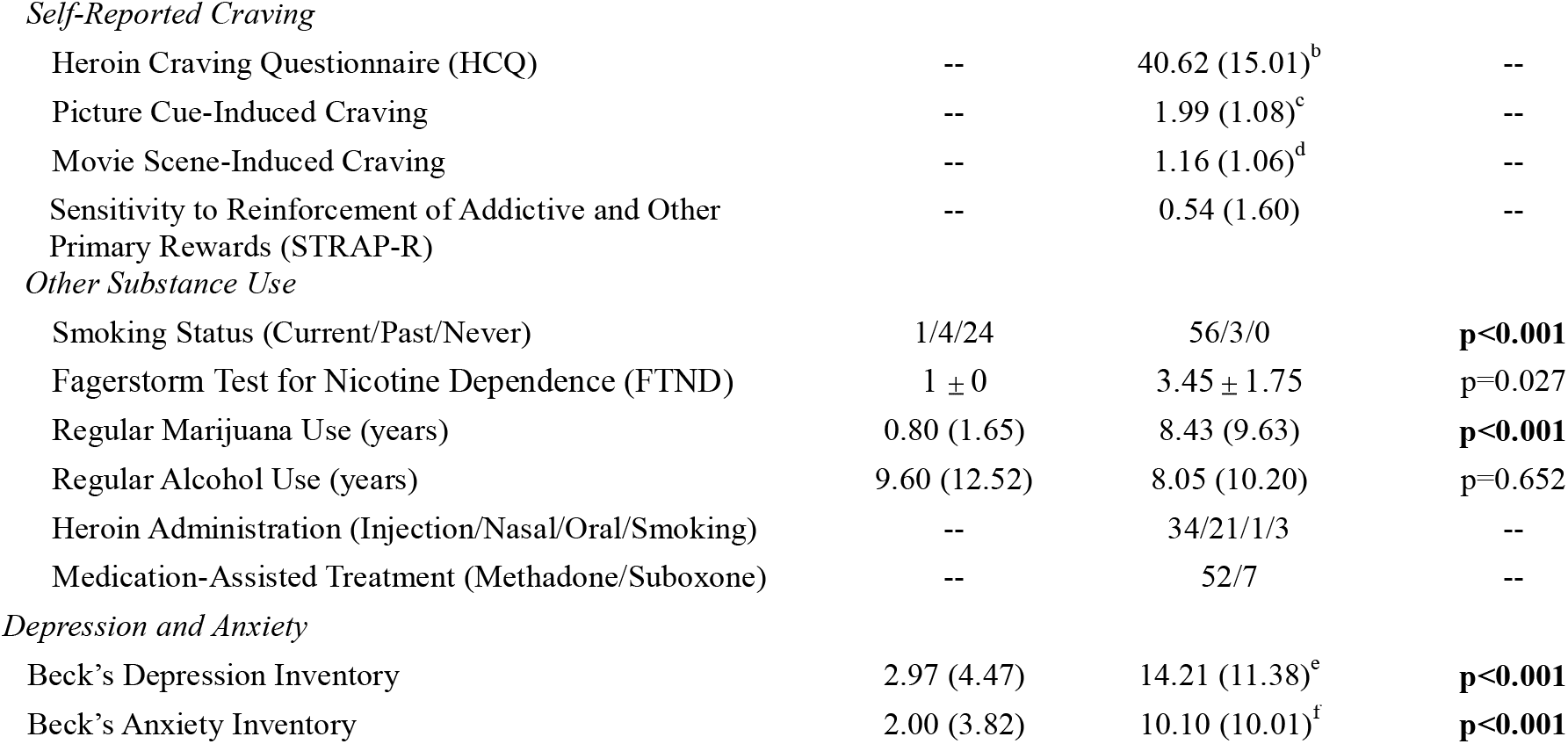
Demographics and drug use variables for all study participants. Values in parentheses denote standard deviation. HC: healthy controls; iOUD: individuals with opioid use disorder. ^a^To assess group differences, Wilcoxon Rank Sum tests were used for all continuous variables except Verbal IQ, where Welch’s t-test was used due to violations of homogeneity of variances. Chi-square tests were used for unordered categorical and binary data. Significant between-group differences, corrected for familywise error (α=.05/12=.0042), are denoted in bold typeface. Handedness was excluded from this correction due to the near parallel distribution between groups. ^b^One missing Heroin Craving Questionnaire score. ^c^One missing cue-induced craving score. ^d^Two missing scene-induced craving scores. ^e^One missing Beck’s Depression Inventory score. ^f^One missing Beck’s Anxiety Inventory score.

|  | HC (N=29) | iOUD (N=59) | sig. test<sup>a</sup> |
| --- | --- | --- | --- |
| *Demographics* |  |  |  |
| Age | 40.70 (10.64) | 41.39 (9.52) | p=0.564 |
| Sex (Male/Female/Other) | 19/10/0 | 44/14/1 | p=0.463 |
| Race (White/Black/Other) | 13/12/4 | 45/5/9 | **p=0.001** |
| Education (years) | 16.22 (2.94) | 12.10 (2.01) | **p<0.001** |
| Verbal IQ | 108.66 (7.30) | 95.54 (10.72) | **p<0.001** |
| Nonverbal IQ | 11.69 (3.06) | 9.76 (3.23) | p=0.005 |
| Handedness (Right/Left) | 22/7 | 46/13 | p=1.000 |
| *Self-Reported Drug-Use Severity* |  |  |  |
| Regular Opioid Use (years) | -- | 11.20 (6.94) | -- |
| Heroin Use Past Month (days) | -- | 0.20 (0.94) | -- |
| Severity Of Dependence Scale (SDS) | -- | 11.66 (3.35) | -- |
| Subjective Opiate Withdrawal Scale (SOWS) | -- | 3.56 (5.51) | -- |
| *Self-Reported Craving* |  |  |  |
| Heroin Craving Questionnaire (HCQ) | -- | 40.62 (15.01)<sup>b</sup> | -- |
| Picture Cue-Induced Craving | -- | 1.99 (1.08)<sup>c</sup> | -- |
| Movie Scene-Induced Craving | -- | 1.16 (1.06)<sup>d</sup> | -- |
| Sensitivity to Reinforcement of Addictive and Other Primary Rewards (STRAP-R) | -- | 0.54 (1.60) | -- |
| *Other Substance Use* |  |  |  |
| Smoking Status (Current/Past/Never) | 1/4/24 | 56/3/0 | **p<0.001** |
| Fagerstorm Test for Nicotine Dependence (FTND) | 1 ± 0 | 3.45 ± 1.75 | p=0.027 |
| Regular Marijuana Use (years) | 0.80 (1.65) | 8.43 (9.63) | **p<0.001** |
| Regular Alcohol Use (years) | 9.60 (12.52) | 8.05 (10.20) | p=0.652 |
| Heroin Administration (Injection/Nasal/Oral/Smoking) | -- | 34/21/1/3 | -- |
| Medication-Assisted Treatment (Methadone/Suboxone) | -- | 52/7 | -- |
| *Depression and Anxiety* |  |  |  |
| Beck's Depression Inventory | 2.97 (4.47) | 14.21 (11.38)<sup>e</sup> | **p<0.001** |
| Beck's Anxiety Inventory | 2.00 (3.82) | 10.10 (10.01)<sup>f</sup> | **p<0.001** |

### 2.2. Drug-Related Cognitive-Behavioral Tasks

Drug choice: We adapted a picture-viewing choice task originally developed for use in cocaine use disorder [21,45,46,72], then adapted for prescription OUD and/or misuse [48,49], methamphetamine use disorder [50], and nicotine smokers [51]. In this study we tailored this picture-viewing task specifically for use in heroin-primary OUD. In brief, choice to view pleasant (e.g., people smiling), unpleasant (e.g., wounds), neutral (e.g., office supplies), and drug pictures (e.g., people using or preparing heroin) was assessed explicitly or probabilistically. In the explicit version, participants made picture-viewing choices (for these four picture categories and blank images as non-stimulus controls) via continued button presses between two simultaneously presented, side-by-side, images. A single press expanded the corresponding image for 0.5 sec, while repeated presses allowed the image to remain expanded for the duration of the trial (5 sec), such that the total responses for each category reflected effort to view these images. In the probabilistic version, participants were prompted to pick one card from four flipped-over decks, pseudorandomly ordered such that each deck contains primarily one category (e.g., drug), while also including pictures from other categories (2 from another category, e.g., pleasant, and 1 from each of the remaining two categories). Choosing a card caused the selected image to fill the screen for 2 sec. Choosing the dominant image category from the same deck a total of eight times ended the run, and unbeknownst to the participant, changed the configuration of the four decks for the next run (similar to [73]), such that subjects needed to relocate their preferred deck. These contingencies were designed to minimize recognition of deck identity while still allowing subjects to establish deck preference [45]. For further task details see [45] and [21].

Drug fluency: Participants were instructed to generate as many drug words (names of people, places, or states of mind related to obtaining, using, or recovering from drugs) as possible in one minute [52,53]. The standard semantic fluency task was administered to measure nondrug fluency, where participants were asked to name as many animals and fruits/vegetables as possible for one minute per category [74].

### 2.3. Attendance of a Follow-Up Visit after Treatment

As a part of the clinical trial associated with this study (NCT04112186), after completing the above tasks (and neuroimaging, reported elsewhere [67,75]), and in parallel to their inpatient standard treatment (described above), the iOUD were randomly assigned to an eight-week group therapy intervention [Mindfulness Oriented Recovery Enhancement [76] (n=28) or a support group (n=30), 1 iOUD withdrawn just prior to randomization; therapy-specific effects will be reported separately in the larger sample that completed this trial]. A follow-up visit consisting of repeating these tasks (and neuroimaging) marked the end of the randomized group treatment, of which the participants were notified during consent with regular reminders throughout the study. Treatment completion was thus defined as attendance of the follow-up session (attended=1, absent=0), which served as an objectively assessed, clinically relevant outcome measure in the regressions below. In the iOUD, reasons for attrition reflected an unwillingness or inability to adhere to the study [e.g., stopped responding to repeated scheduling efforts (n=10), unwilling to continue (n=4), withdrawn due to study non-compliance (n=2)], lending support to the idea that this follow-up attendance is a valid measure of treatment adherence.

The 29 HC subjects were studied at similar time intervals, all attending both the baseline and follow-up sessions, yielding significant group differences in follow-up attendance rates [*X*^2^ (1, N=88)=7.88, *p*=0.005]. The behavioral tasks were conducted twice, at baseline and follow-up, an average of 17 weeks apart (119±56 days). Group differences in the days between sessions did not reach significance (iOUD: 108±36; HC: 136±76; *Z*=-1.35, *p*=0.178).

### 2.4. Statistical Analysis

#### Choice Behavior and Fluency

For the explicit and probabilistic choice tasks, the total response count and total image selections, respectively, per picture category across all trials yielded category-specific choice. The a priori selected drug>pleasant contrast estimated drug-biased choice (controlling for a competing nondrug reward within-subjects). Drug and non-drug fluency scores were calculated by summing correct responses per minute, excluding repetitions and errors (i.e., a sum for the 1-min drug category and an average across the two 1-min nondrug categories). The drug>nondrug contrast represented drug-biased fluency.

Three mixed analyses of variance (ANOVAs) were conducted for the primary behavioral variables at baseline: 1) for explicit choice, a 2 (group: iOUD, HC) × 5 (cue type: drug, pleasant, unpleasant, neutral, blank) ANOVA; 2) for probabilistic choice, a 2 (group: iOUD, HC) × 4 (cue type: drug, pleasant, unpleasant, neutral) ANOVA (2 iOUD and 1 HC missing); 3) for fluency, a 2 (group: iOUD, HC) × 2 (cue type: drug, nondrug) ANOVA (1 iOUD missing). Across all three measures, longitudinal effects were estimated using mixed ANOVAs with an additional factor for session (baseline, follow-up; see supplement); correlations between baseline and follow-up behavior were also conducted across all three measures (see supplement).

Significant interactions (*p*=.05) were followed by paired and independent parametric t-tests. Non-parametric t-tests (Wilcoxon tests) were used in cases where assumptions of normality were violated (via the Shapiro-Wilk test); Welch’s t-tests were used in cases where assumptions of homogeneity of variances were violated (via the Bartlett test).

To assess the putative contribution of measures showing significant group differences (Table 1), Pearson correlations were conducted with the primary baseline behavioral measures. The measures showing significant associations were entered in subsequent ANCOVAs (see supplement).

The following analyses pertain to the iOUD group only.

#### Factor Analysis

Given our iOUD sample size and thereby the need for dimensionality reduction, we set to identify the variables most representative of our hypothesized regressor categories, conducting a factor analysis using select demographics measures, subjective drug-use severity and craving measures (from Table 1), and the objective drug-biased cognitive-behavioral measures; that is, we aimed to reduce the number of predictor variables to 4 (ratio variable: sample size=1:15). Using a standard factor analysis approach, unordered categorical (sex, race, smoking status, route of heroin administration) and binary (handedness and stabilizing medication) data were omitted. Years of regular marijuana, alcohol use, and FTND were removed as the intended focus for the self-reported measures was their primary drug. Heroin use in the past month was excluded due to its lack of variance in this abstinent population. Anxiety and depression were excluded as they did not represent our pre-selected categories. The raw values of the variables selected from this factor analysis (those with the highest loadings on each factor) were to become the regressors in the subsequent analyses.

#### Hierarchical Logistic Regression

A hierarchical logistic regression using the categorical treatment completion (i.e., attendance of the follow-up session) as the dependent variable and a stepwise inclusion of the regressors that emerged from the factor analysis was performed. We intended to assess the unique contributions of our select objective drug-biased measures against subjective and demographics measures in predicting clinical outcome. Specifically, the first model included only the demographics variable (“demographics model”), which was compared to the second model comprising the demographics and subjective severity measures (“subjective severity model”). The second model was then compared to the third model, which included the subjective craving measure and the second model’s regressors (“subjective craving model”). The third model was then compared to the fourth model, which included the objective drug bias marker and the third model’s regressors (“objective model”), effectively comparing the objective and subjective measures in explaining treatment adherence, controlling for demographics. Likelihood ratio tests were conducted for the nested model comparisons and the model summaries (via null model comparisons).

## 3. RESULTS

The iOUD and HC were comparable in age, sex, nonverbal IQ, handedness, and years of regular alcohol use. There were significant group differences in race (white and other, iOUD>HC; black HC>iOUD), years of education and verbal IQ (HC>iOUD), depression and anxiety symptoms (iOUD>HC), smoking status (current, iOUD>HC; past and never, HC>iOUD) and years of regular marijuana use (iOUD>HC); *ps*≤0.005 (family-wise corrected across 11 comparisons), see Table 1. These group differences did not contribute to the ANOVA results reported below (see supplement). There were no significant differences in these variables between the follow-up attendance iOUD subgroups (see Table S1 in supplement), hence no additional measures were controlled for in the regression models.

### 3.1. Fluency and Choice Behavior

Explicit choice results revealed no main effect of group [*F*(1,86)=0.72, *p*=0.399], a main effect of cue type [pleasant>neutral>blank=unpleasant>drug; *F*(4,344)=76.61, *p*<0.001], and a group × cue type interaction [*F*(4,344)=10.75, *p*<0.001]. This interaction was driven by more drug and unpleasant image choice (*Z*>2.09, *p*<0.037) and less pleasant, neutral, and blank image choice [*t*(86)>2.58 or Z>2.43, *p*<0.015] in the iOUD compared to HC; within groups, drug image choice was lower than choice for all other categories (iOUD: Z>3.00, *p*<0.003; HC: Z>2.07, *p*<0.039) (Figure 1A).

**Figure 1.**
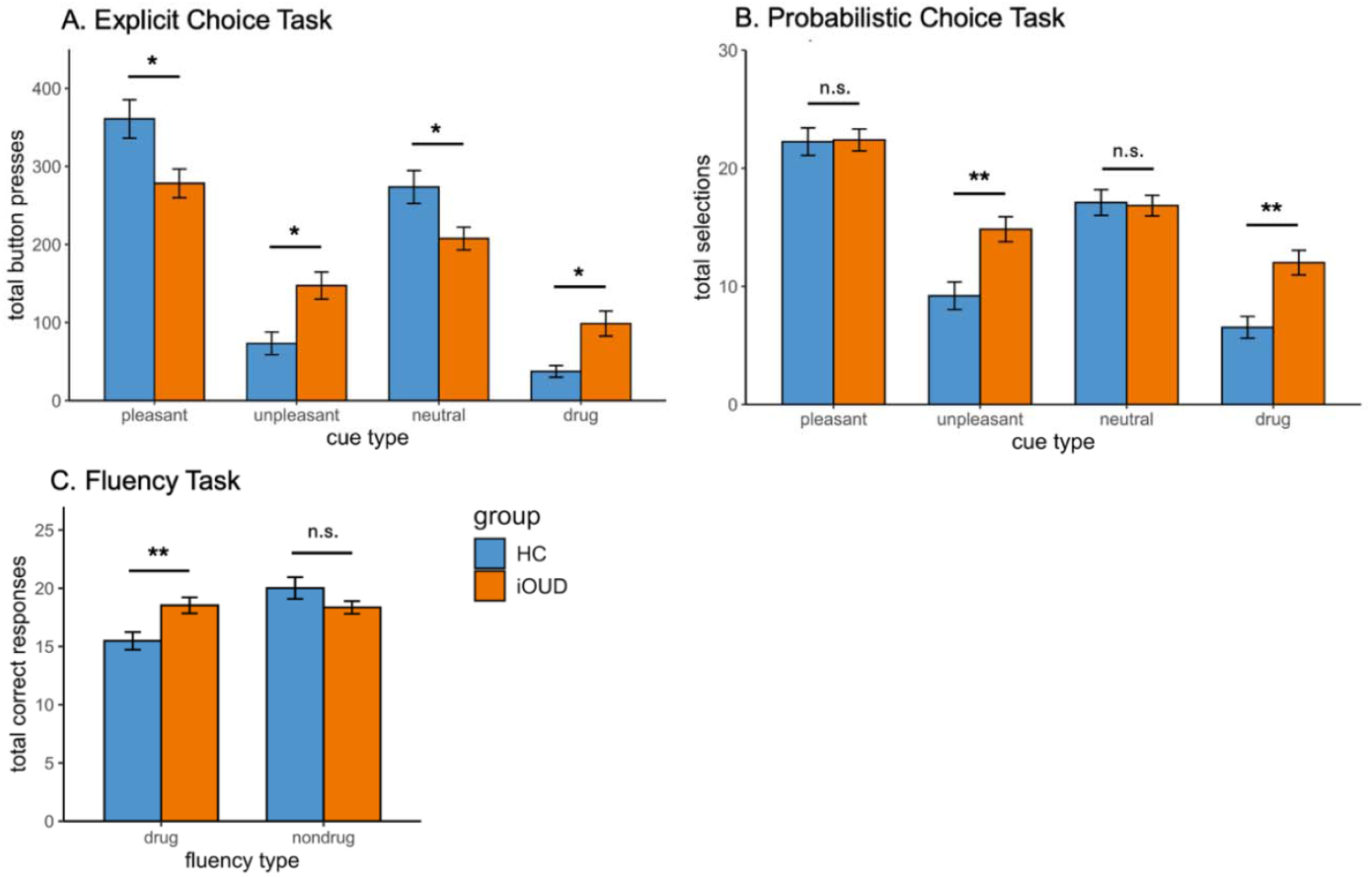
Choice and fluency behavior. HC: healthy controls; iOUD: individuals with opioid use disorder. iOUD and HC participants’ (A) explicit choice behavior, indicating a bias in iOUD towards drug (p=0.036) and unpleasant (p=0.014) cue types and a bias in HC towards pleasant (p=0.010) and neutral (p=0.011) cue types, (B) probabilistic choice behavior, indicating a bias in iOUD towards drug (p=0.002) and unpleasant (p=0.002) cue types, with no group differences in pleasant (p=0.840) and neutral (p=0.857) cue types, and (C) fluency behavior, indicating drug-biased fluency in iOUD (p=0.008) with no group differences in nondrug fluency (p=0.103). Non-stimulus cues are not visualized. Error bars indicate SEM. Asterisks reflect significant group differences (*p<0.05 and ** p<0.01).

Probabilistic choice results revealed a main effect of group [iOUD>HC; *F*(1,83)=8.10, *p*=0.006] and cue type [pleasant>neutral>unpleasant>drug; *F*(3,249)=61.79, *p*<0.001], and a group × cue type interaction [*F*(3,249)=4.92, *p*=0.002]. This interaction was driven by more drug and unpleasant [*Z*>3.12, *p*<0.002] image choice in iOUD compared to HC, with no group differences for pleasant or neutral images [*t*(83)=0.18 or Z=0.20, *p*<0.857]; within groups, selection of drug images was lower than choice for all other categories [iOUD: *Z*>3.79, *p*<0.001; HC: *Z*>3.30, *p*<0.001] (Figure 1B).

Fluency results revealed no main effect of group [*F*(1,85)=0.61, *p*=0.439], a main effect of cue type [nondrug>drug; *F*(1,85)=14.24, *p*<0.001], and a group × cue type interaction [*F*(1,85)=16.70, *p*<0.001], revealing more drug word generation by iOUD compared to HC [Z=2.64, *p*=0.008] with no difference in nondrug words [*t*(85)=1.65, *p*=0.103]; within groups, the HC generated more nondrug than drug words [*t*(28)=5.00, *p*<0.001], not observed in iOUD [Z=0.01, *p*=0.990] (Figure 1C).

There were no session effects nor longitudinal 2- or 3-way interaction effects detected for any of the tasks (*p*s>0.113, see supplement); within-subject correlations revealed associations between baseline and follow-up behavior (*p*s<0.010; see Figure S2 in supplement).

#### 3.2. Factor Analysis

The factor analysis yielded four factors where the variables with the highest loadings were consistent with our predetermined categories: education (years) (demographics; λ=0.52), regular opioid use in years (subjective severity measure; λ=0.95); picture cue-induced craving (subjective craving measure; λ=0.93); and drug>pleasant explicit choice (objective marker; λ=0.99) (Table 2).

**Table 2.**
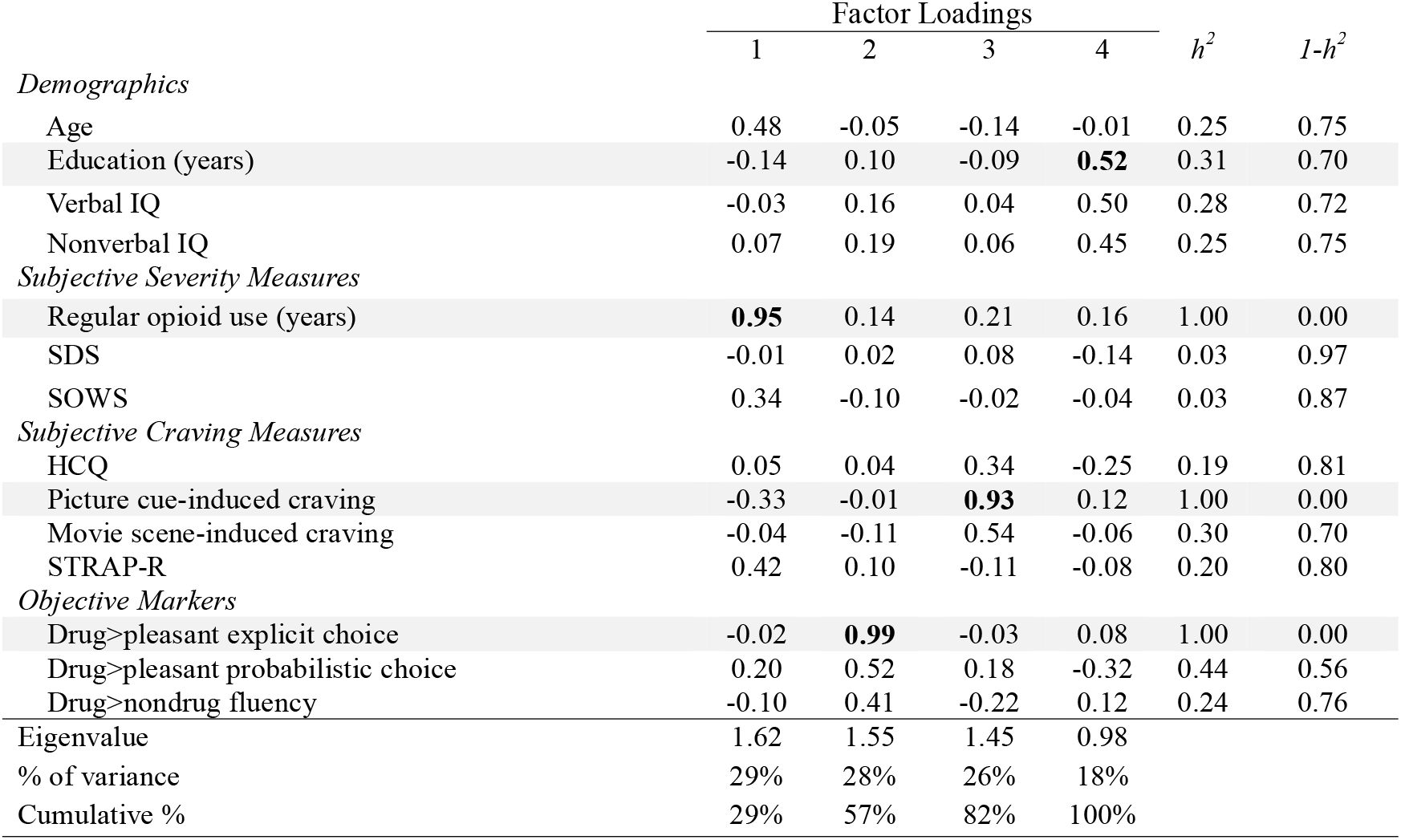
Factor analysis for dimension reduction. Factor loadings, communality (*h*^*2*^), uniqueness (*1-h*^*2*^), eigenvalues, percentages of variance and cumulative percentages of variance are shown. The extraction method used was maximum likelihood estimation with a varimax rotation method. Conducted in 52 individuals with opioid use disorder (iOUD) with complete data for all variables. SDS=Severity of Dependence Scale; SOWS=Subjective Opiate Withdrawal Scale; HCQ=Heroin Craving Questionnaire; STRAP-R=Sensitivity to Reinforcement of Addictive and Other Primary Rewards.

#### 3.3. Hierarchical Logistic Regression

Hierarchical regression results revealed that the first model was not significant (*R*^*2*^=0.06, *p*=0.097), such that demographics (years of education) alone were not significantly associated with treatment completion (*β*=0.58, *p*=0.122). The second model including the subjective severity measure (years of regular opioid use) was also not significant (*R*^*2*^=0.08, *p*=0.191) and did not perform significantly better compared to demographics alone (_Δ_*R*^*2*^=0.014, *p*=0.457). Similarly, the third model with subjective craving (picture cue-induced craving) was not significant (*R*^*2*^=0.08, p=0.312) and did not perform significantly better compared to the subjective severity model (_Δ_*R*^*2*^=0.003, p=0.611). However, adding the objective marker (drug>pleasant explicit choice) significantly increased the variability in the likelihood of treatment completion explained by the model, increasing the pseudo-R^2^ by 10.2% (*p*=0.027). In this final model, the objective measure was associated with the clinical outcome such that the higher the baseline drug bias, the lower the likelihood of treatment completion (*β* =-0.75, *p*=0.036), not seen for the demographic, subjective severity and craving measures (*ps>*0.061). In the final model, 18% of the variance in the likelihood of treatment completion was associated with the model regressors (p=0.075; Table 3). In a separate logistic regression, a model including only the drug-biased behavior measure (controlling for demographics) reached significance compared to the null (AIC=59.9, pseudo-R^2^=0.13, p=0.035), while independently regressing regular opioid use and picture cue-induced craving (each controlling for demographics) did not (*ps*>0.191).

**Table 3.** Hierarchical regression analysis of predictors of treatment completion. Hierarchical logistic regression results are shown for the 52 individuals with opioid use disorder (iOUD) included in the factor analysis. For each logistic regression, the standardized coefficient estimates (*β*), standard errors (SE), and p-values are displayed for each predictor variable. Summary statistics (AIC, pseudo-R^2^, and p-values) are displayed for each model vs. the null model. Increases in R^2^ values and p-values are displayed for each step’s model comparison. P-values below 0.05 are denoted in bold typeface.

| | $\beta$ | SE | p-value | Model summary | | | Model Comparison | | |
| --- | --- | --- | --- | --- | --- | --- | --- | --- | --- |
| | | | | AIC | R<sup>2</sup> | p-value | Test | $\Delta R^2$ | p-value |
| *1. Demographics Model* |  |  |  | 61.8 | 0.06 | 0.097 | -- | -- | -- |
| Education (years) | 0.58 | 0.18 | 0.122 |  |  |  |  |  |  |
| *2. Subjective Severity Model* |  |  |  | 63.3 | 0.08 | 0.191 | 1 vs. 2 | 0.014 | 0.457 |
| Education (years) | 0.60 | 0.19 | 0.113 |  |  |  |  |  |  |
| Regular opioid use (years) | 0.26 | 0.05 | 0.470 |  |  |  |  |  |  |
| *3. Subjective Craving Model* |  |  |  | 65.0 | 0.08 | 0.312 | 2 vs. 3 | 0.003 | 0.611 |
| Education (years) | 0.60 | 0.18 | 0.112 |  |  |  |  |  |  |
| Regular opioid use (years) | 0.24 | 0.05 | 0.502 |  |  |  |  |  |  |
| Picture cue-induced craving | -0.16 | 0.30 | 0.608 |  |  |  |  |  |  |
| *4. Objective Model* |  |  |  | 62.1 | 0.18 | 0.075 | 3 vs. 4 | 0.102 | **0.027** |
| Education (years) | 0.77 | 0.20 | 0.061 |  |  |  |  |  |  |
| Regular opioid use (years) | 0.47 | 0.06 | 0.290 |  |  |  |  |  |  |
| Picture cue-induced craving | -0.21 | 0.31 | 0.533 |  |  |  |  |  |  |
| Drug>pleasant explicit choice | -0.75 | 0.00 | **0.036** |  |  |  |  |  |  |

## 4. DISCUSSION

Clinical research in addiction traditionally relies on subjective reports (questionnaires, interviews) of drug-use severity to characterize treatment endpoints, with limited success [77–79]. Here, in inpatient iOUD we show that objective cognitive-behavioral markers of drug cue reactivity, assessed with well-validated tasks that tap into a core addiction symptom (as postulated by the iRISA model), outperformed their subjective (and demographic) counterparts in informing prospective treatment adherence. First, in heroin-primary OUD we replicated findings previously demonstrated mostly in cocaine addiction [45,52] by showing drug-biased choice and fluency behavior. Compared to HC, iOUD selected more drug (and unpleasant) images and generated more drug words, supporting generalizability of these measures as markers of drug bias in addiction; measures that differed between the groups (Table 1) did not contribute to these results (see supplement). Following dimensionality reduction, we then used a hierarchical regression model to show that only the objective drug bias marker (drug>pleasant explicit choice) significantly accounted for the variance in treatment completion in the iOUD group, outperforming the subjective markers (years of regular opioid use and cue-induced craving), while controlling for demographics (years of education). These novel results illustrate that disease-targeted objective behavioral measures may serve as valuable markers of addiction severity to inform prospective treatment adherence in iOUD.

The heightened drug-biased behavior we report extends to treatment-seeking iOUD previous findings mostly in cocaine addiction obtained with both the picture choice [45] and drug fluency [52] tasks. This drug-biased picture choice and verbal fluency are consistent with biased attentional allocation (via the dot probe pictorial task) towards drug stimuli in methadone-maintained outpatients with heroin addiction [80] and opioid-dependent pain patients [42,81]. Together, this drug bias is consistent with an enhanced salience (encompassing attentional bias, reactivity and subjective valence) attributed to drug over nondrug cues, with a concomitant inhibitory control dysfunction, in people with addiction; deficits that the iRISA model identifies as rooted in the mesencephalic prefrontal cortico-limbic processes [53,72] that perpetuate the cycle of relapse in people with addiction [7,8]. Unlike previous reports in cocaine addiction [45], the choice task results also revealed a bias towards unpleasant images. This finding aligns with heightened emotional responses to unpleasant>pleasant imagery in current opioid users [82], and biased attention to negative>positive facial expressions in abstinent heroin users [83]. Overall, results may reflect frequent exposure to unpleasant stimuli in the living environment and/or dysregulated affective processing in iOUD [82,83], consistent with their higher rates of depression and anxiety [84–86], as also seen here. Targeted investigations of this negative choice bias are needed to better understand its underlying neural substrates and clinical relevance in iOUD.

For the first time in drug addiction, we show that compared to subjective self-report measures (and controlling for demographics), objective behavioral measures of drug bias (drug>pleasant picture choice) can better account for future treatment adherence, also objectively measured (attendance of a treatment follow-up visit). Indeed the self-reported measures (regular opioid use and cue-induced craving) did not significantly predict clinical outcome, consistent with prior reports of these measures’ weak or null relationships with outcomes such as relapse in cocaine and alcohol dependence ([87]; but see also [13,88,89]) and treatment dropout in cocaine [90,91] and other substance use disorders ([92]; but see also [93]). In contrast, the significant contribution of objective behavioral measures to outcome aligns with previous work showing that drug-biased picture choice prospectively predicted 6-months self-reported drug use in cocaine addiction [46] and an 8-week follow-up opioid misuse severity (via the Addictions Behavior Checklist) in chronic pain patients [48]. Other picture-viewing paradigms that model approach and avoidance tendencies towards drug images have prospectively predicted 6-months cannabis use (via structured interviews/questionnaires) in heavy marijuana users [94]. The predictive value of these drug-biased behavioral measures may stem from their objective probing of conditioned (automatic) responses to drug cues (e.g., attentional bias, physiological arousal) [95,96] shown to contribute to future relapse [97]. Another contributing factor may be their increased reliance on higher-order cognitive functions (e.g., working-memory, decision-making, recall) that are commonly impaired in addiction [7] as associated with poor treatment outcomes [36,98–100]. We postulate that the modulation of these cognitive functions by a drug-related context confers the greatest predictive power, exemplifying the subversion of these functions towards the processing of drug and drug-related cues at the expense of their nondrug counterparts in people with addiction.

Several limitations should be noted when interpreting these results. First, while the measures that showed group differences (see Table 1) did not contribute to our results, samples more closely matched along demographics, smoking status, and marijuana use could better accommodate the evaluation of their contribution. A larger sample size could also allow examining a larger number of subjective and objective measures, as well as potential sex differences, in our results. Additionally, the treatment completion measure chosen reflects the therapy provided in addition to standard inpatient treatment, not allowing insight into adherence to the latter. Future studies should investigate whether these objective drug bias markers can explain additional clinical endpoints as assessed in other treatment contexts (e.g., outpatient medication-assisted or inpatient treatment, varying treatment durations).

There is an urgent need to develop and test measures that can be used to characterize drug-use severity and inform drug treatment outcomes to counteract the treatment-resistant nature of addiction. Here we show that choice and fluency tasks can serve as reliable markers of drug-biased behavior in iOUD. We also report that these objective markers of drug bias outperformed traditionally employed subjective markers (self-reports of duration of use and cue-induced craving, controlling for demographic confounds) in predicting future treatment adherence. Assessing a core deficit in drug addiction (i.e., attribution of salience and other cognitive resources towards drug over alternative cues) while circumventing the limitations of self-report and non-varying categorical measures (e.g., drug toxicology), these objective cognitive-behavioral measures of drug bias present viable alternatives and/or supplements to the most common outcome measures utilized in clinical trials and other drug addiction research. Their use may lead to improved treatment models, allowing for early risk identification and the deployment of effective prevention efforts.

## ACKNOWLEDGEMENTS

We would like to thank Yuefeng Huang, Pierre-Olivier Gaudreault, Sarah King, Pias Malaker, Pazia Miller, Amelia Brackett, Gabriela Hoberman, Devarshi Vasa, Defne Ekin, Chloe Wong, Lucy Bao, Rachel Drury, and Maggie Boros for their assistance in data collection.

## AUTHOR CONTRIBUTIONS

NM: Conceptualization, data collection, analysis, writing the initial draft; AC: Supervision, conceptualization, data collection, revision of manuscript; GK: Conceptualization, data collection; NA: Supervision, revision of manuscript; RG: Funding acquisition, supervision, conceptualization, revision of manuscript.

## FUNDING

This work was supported by R01AT010627 to RZG.

## COMPETING INTERESTS

The authors have nothing to disclose.

## DATA AVAILABILITY STATEMENT

The data that support these results are available upon reasonable request from the corresponding author, as participants are a vulnerable population.

## Notes

### Competing Interest Statement

The authors have declared no competing interest.

### Clinical Trial

NCT04112186

### Funding Statement

This work was supported by NCCIH grant R01AT010627 to Dr. Goldstein.

### Author Declarations

The Icahn School of Medicine at Mount Sinai institutional review board approved study procedures, and all participants provided written informed consent.

