## Supplement for "Moving beyond self-report in characterizing drug addiction: Using drug-biased behavior to prospectively inform treatment adherence in opioid use disorder"

*Eligibility Criteria*

All participants met the following inclusion criteria: 1) Ability to understand and give informed consent; and 2) 18-64 years of age. All iOUD further met the following inclusion criteria: 1) Diagnostic and Statistical Manual of Mental Disorders (DSM-5) diagnosis of OUD with heroin as the primary drug of choice; and 2) stabilized on medication-assisted treatment (i.e., methadone or suboxone).

Participants were excluded from the study if they met any of the following criteria: 1) DSM-5 diagnosis for schizophrenia or developmental disorder (e.g., autism); 2) head trauma with loss of consciousness (>30 min); 3) history of neurological disease of central origin including seizures; 4) cardiovascular disease including high blood pressure and/or other medical conditions, including metabolic, endocrinological, oncological or autoimmune diseases, and infectious diseases common in iOUD (including Hepatitis B and C or HIV/AIDS); 5) metal implants or other MR contraindications (e.g., claustrophobia); and 6) women who were pregnant or lactating. To recruit a participant sample most representative of OUD in the real world, iOUD were not excluded for a DSM-5 diagnosis of a substance use disorder other than opiates, as long as opiates were the primary drug of choice and/or reason for treatment. Healthy controls were excluded if they met DSM-5 criteria for a substance use disorder; testing positive for drugs was also exclusionary.

*Psychiatric Diagnoses and Comorbid Substance Use Disorders*

In the OUD, other psychiatric diagnoses included depressive episode disorder (n=30; 4 current), suicidality/suicidal behavior (n=5; 0 current), panic disorder (n=5; 3 current), agoraphobia (n=2; 1 current), obsessive compulsive disorder (n=1; 1 current), post-traumatic stress disorder (n=12; 6 current), binge eating disorder (n=1; 1 current), and generalized anxiety disorder (n=2; 2 current). Other substance use disorders included cocaine (n=20; 3 in sustained remission, 9 in early remission, 8 current), sedatives (n=10; 2 in sustained remission, 4 in early remission, 4 current), cannabis (n=3; all in early remission), non-crack/cocaine stimulants (n=1; in early remission), and polysubstance use disorder (n=1; in sustained remission). In the HC group, psychiatric diagnoses included depressive episode disorder (n=3; 0 current) and suicidality (n=1; 0 current).

*Demographics and Drug Use by Follow-Up Attendance*

The follow-up attendance groups did not differ in demographics measures (*ps*>0.043), self-reported drug use severity measures (ps>0.122), self-reported craving measures (*ps*>0.034), other substance use variables (*ps*>0.027), or depression and anxiety scores (ps>0.922) after correcting for familywise error (α=.05/22=.002). See Table S1 for details on these measures between the follow-up attendance iOUD groups.

Table S1. Demographics and drug use by follow-up attendance.

|  |  | Attended (n=38)^a^ | Absent (n=14)^a^ | sig. test^b^ |
| --- | --- | --- | --- | --- |
|  | *Demographics* |  |  |  |
|  | Age | 42.50 (10.30) | 37.39 (7.96) | p=0.083 |
|  | Sex (Male/Female/Other) | 28/9/1 | 12/2/0 | p=0.610 |
|  | Race (White/Black/Other) | 27/5/6 | 13/0/1 | p=0.218 |
|  | Education (years) | 12.37 (1.99) | 11.36 (2.10) | p=0.043 |
|  | Verbal IQ | 96.18 (9.75) | 94.00 (11.10) | p=0.493 |
|  | Nonverbal IQ | 9.79 (2.93) | 8.71 (4.01) | p=0.295 |
|  | Handedness (Right/Left) | 30/8 | 12/2 | p=0.879 |
|  | *Self-Reported Drug Use Severity* |  |  |  |
|  | Regular Opioid Use (years) | 11.38 (7.56) | 10.04 (5.77) | p=0.733 |
|  | Heroin Use Past Month (days) | 0.32 (1.16) | 0.00 (0.00) | p=0.122 |
|  | Severity Of Dependence Scale (SDS) | 11.24 (3.51) | 12.14 (3.18) | p=0.358 |
|  | Subjective Opiate Withdrawal Scale (SOWS) | 3.39 (5.53) | 3.00 (3.31) | p=0.871 |
|  | *Self-Reported Craving* |  |  |  |
|  | Heroin Craving Questionnaire (HCQ) | 38.79 (13.54) | 48.93 (18.14) | p=0.034 |
|  | Picture Cue-Induced Craving | 1.98 (1.07) | 2.17 (1.07) | p=0.419 |
|  | Movie Scene-Induced Craving | 1.18 (1.09) | 1.08 (0.70) | p=0.820 |
|  | Sensitivity to Reinforcement of Addictive and Other  Primary Rewards (STRAP-R) | 0.34 (1.42) | 1.14 (1.92) | p=0.289 |
|  | *Other Substance Use* |  |  |  |
|  | Smoking Status (Current/Past/Never) | 37/1/0 | 13/1/0 | p=1.000^c^ |
|  | Fagerstorm Test For Nicotine Dependence (FTND) | 3.08 (1.89)^d^ | 4.38 (1.33)^e^ | p=0.027 |
|  | Regular Marijuana Use (years) | 8.18 (9.81) | 8.75 (8.78) | p=0.668 |
|  | Regular Alcohol Use (years) | 7.95 (10.36) | 6.82 (7.41) | p=0.890 |
|  | Heroin Administration (Injection/Nasal/Oral/Smoking) | 18/17/1/2 | 10/3/0/1 | p=0.456 |
|  | Medication-Assisted Treatment (Methadone/Suboxone) | 34/4 | 11/3 | p=0.573 |
| *Depression and Anxiety* | |  |  |  |
|  | Beck’s Depression Inventory (BDI) | 14.45 (11.58) | 15.85 (13.32)^f^ | p=0.965 |
|  | Beck’s Anxiety Inventory (BAI) | 10.34 (8.80) | 12.46 (14.16)^g^ | p=0.922 |
|  | ^a^Differences between follow-up attendance groups across Table 1 measures are displayed for the 52 iOUD with complete data in the factor analysis, the subset then used for the regression analyses.  ^b^To assess group differences across the continuous variables displayed in S1, t-tests were used for normally distributed variables, and Welch’s t-test and Wilcoxon rank sum tests were used when assumptions of homogeneity of variance and normality were violated, respectively. Chi-square tests were used for unordered categorical and binary data comparisons. Significant between-group differences were corrected for familywise error (α=.05/22=.002), with smoking status excluded due to a near parallel distribution.  ^c^Smoking status “Never” was excluded from Chi-square test as it had no occurrences in either group.  ^d,e^One missing FTND score.  ^f^One missing BDI score.  ^g^One missing BAI score. | | | |

*Examining Recovery in Drug-Biased Behavior*

Longitudinal analyses of explicit choice (39 iOUD, 25 HC) revealed no main effect of group [*F*(1,62)=0.11, *p=*0.737], a significant main effect of cue type [pleasant>neutral>blank>negative>drug; *F*(4,248) = 86.21, *p*<0.001], no main effect of session [*F*(1,62)=0.03, *p=*0.856], a significant group*cue-type interaction [*F*(4,248)=8.02, *p*<0.001], no group*session [*F*(1,62)=0.53, *p*=0.468] or cue-type*session [*F*(4,248)=1.25, *p*=0.292] interaction, and no 3-way interaction [*F*(4,248)=0.85, *p=*0.495]. For probabilistic choice (38 iOUD, 24 HC), there was a main effect of group [iHUD>HC; *F*(1,60)=7.54, *p*=0.008] and cue-type [pleasant>neutral>unpleasant>drug; *F*(3,180)=73.18, *p<*0.001] but not session [*F*(1,60)=0.46, *p*=0.500]*,* a significant group*cue-type interaction [*F*(3,180)=4.56, *p*=0.004], no group*session [*F*(1,60)=0.79, *p*=0.379] or cue-type*session [*F*(3,180)=0.55, *p*=0.650] interaction, and no 3-way interaction [*F*(3,180)=0.77, *p=*0.510]. For fluency (32 iOUD, 26 HC), there was no main effect of group [*F*(1,56)=2.08, *p=*0.155] or session [*F*(1,56)=0.28, *p=*0.602], a significant main effect of fluency type [nondrug>drug; *F*(1,56) = 26.78, *p<*0.001], a significant group*fluency-type interaction [*F*(1,56)=17.06, *p*<0.001], no group*session [*F*(1,56)=1.70, *p*=0.197] or session*fluency-type [*F*(1,56)=2.59, *p*=0.113] interaction, and no 3-way interaction [*F*(1,56) = 1.16, *p=*0.286] (see Figure S1). These results are largely consistent with those reported in the main text, where behavior showed no change between both administrations.

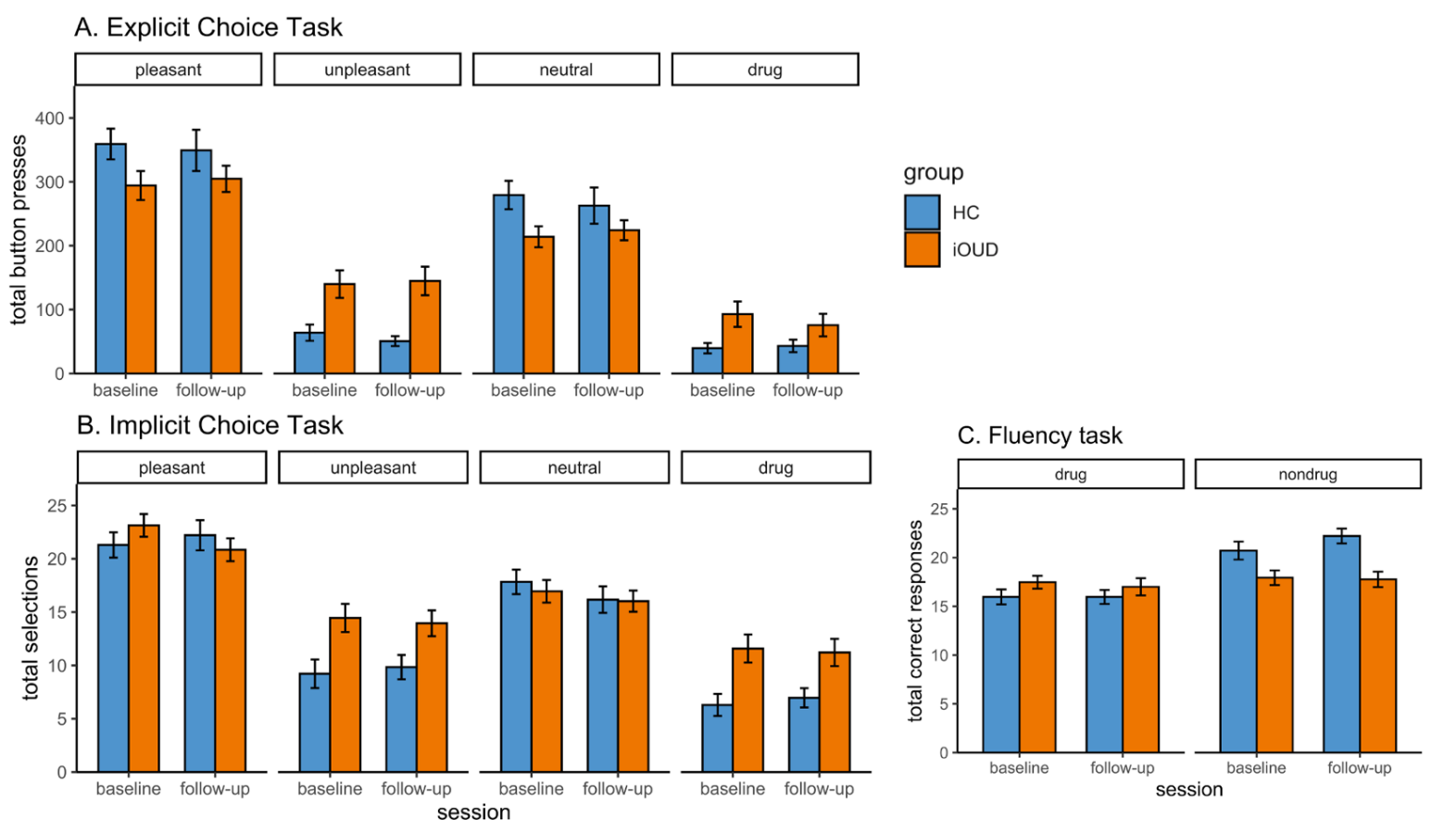
Figure S1. Longitudinal results in choice and fluency behavior.

Individuals with opioid use disorder (iOUD) and healthy control (HC) participants’ (A) explicit choice behavior, (B) probabilistic choice behavior, and C) fluency behavior revealing no reductions in drug-biased behaviors between baseline and follow-up in the subsets of participants with complete task data. Non-stimulus cues are not visualized. Error bars indicate SEM.

To further investigate task stability, we inspected correlations between baseline and follow-up performance within-subjects across tasks. Results revealed moderate to strong correlations (*r>*0.41, *p*<0.010), illustrating high test-retest reliability.

Figure S2. Intra-task correlations in choice and fluency behavior.

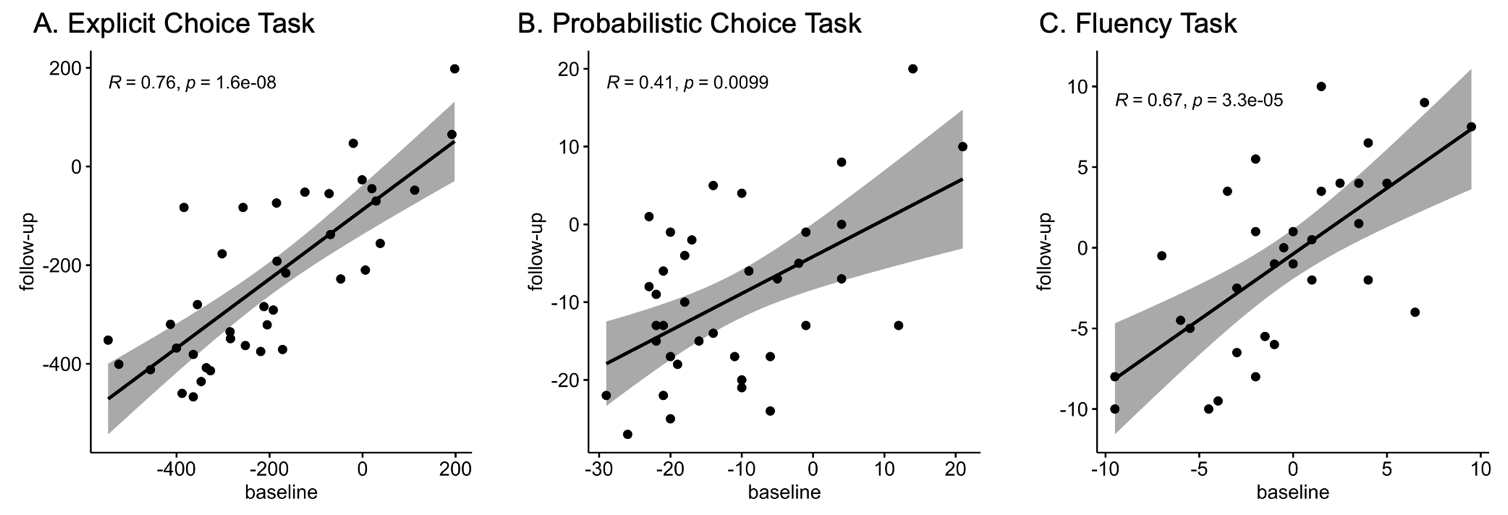

Within-subject, intra-task correlations for (A) explicit choice (B) implicit choice and (C) fluency show positive correlations between baseline and follow-up behavior in the individuals with opioid use disorder (iOUD) with data at both timepoints.

*Examining Potential Covariates in Baseline Analyses*

To examine potential contribution to our results, for variables that showed significant group differences (see Table 1), we performed Pearson correlations with our main behavioral measures (drug>pleasant explicit and probabilistic choice, and drug>non-drug fluency). Smoking status was excluded due to its near parallel distribution between groups. Race was dummy coded for white, black and other. The Table 1 measures that differed between the groups and showed significant associations with the behavioral measures [after correction for familywise error, α=.05/(6*3)=.003], were individually controlled for using group × cue-/fluency-type ANCOVAs to assess whether the group*cue-/fluency-type interaction effects survived.

Correlations with explicit choice revealed that, while race, education, verbal IQ, and regular marijuana use were not associated with drug>pleasant choice (*p*s>0.075), anxiety and depression scores were (*p*<0.002). However, controlling for depression and anxiety in the model did not affect the results (*p*s<0.001). Correlations with probabilistic choice revealed that none of the Table 1 measures were significantly associated with drug>pleasant choice (*p*s>0.014). Correlations with fluency revealed that, while race, education, verbal IQ, regular marijuana use, and anxiety scores were not associated with drug>non-drug fluency (*p*s>0.010), depression scores were (*p*=0.001). However, controlling for depression in the model did not affect the results (*p*=0.008).
